# Hyperacute Prediction of Targeted Temperature Management Outcome After Cardiac Arrest

**DOI:** 10.1101/2023.06.12.23291152

**Authors:** Jocelyn Hsu, Han Kim, Kirby Gong, Tej D. Azad, Robert D. Stevens

## Abstract

**Introduction:** Targeted temperature management (TTM) has been associated with greater likelihood of neurological recovery among comatose survivors of cardiac arrest. However, the efficacy of TTM is not consistently observed, possibly due to heterogeneity of therapeutic response. The aim of this study is to determine if models leveraging multi-modal data available in the first 12 hours after ICU admission (hyperacute phase) can predict short-term outcome after TTM.

**Methods:** Adult patients receiving TTM after cardiac arrest were selected from a multicenter ICU database. Predictive features were extracted from clinical, physiologic, and laboratory data available in the hyperacute phase. Primary endpoints were survival and favorable neurological outcome, determined as the ability to follow commands (motor Glasgow Coma Scale [mGCS] of 6) upon discharge. Three machine learning (ML) algorithms were trained: generalized linear models (GLM), random forest (RF), and gradient boosting (XG). Models with optimal features from forward selection were 10-fold cross-validated and resampled 10 times.

**Results:** Data were available on 310 cardiac arrest patients who received TTM, of whom 183 survived and 123 had favorable neurological outcome. The GLM performed best, with an area under the receiver operating characteristic curve (AUROC) of 0.86 ± 0.04, sensitivity 0.75 ± 0.09, and specificity 0.77 ± 0.07 for the prediction of survival and an AUROC of 0.85 ± 0.03, sensitivity 0.71 ± 0.10, and specificity 0.80 ± 0.12 for the prediction of favorable neurological outcome. Features most predictive of both endpoints included lower serum chloride concentration, higher serum pH, and greater neutrophil counts.

**Conclusion:** In patients receiving TTM after cardiac arrest, short-term outcomes can be accurately discriminated using ML applied to data routinely collected in the first 12 hours after ICU admission. With validation, hyperacute prediction could enable personalized approach to clinical decision-making in the post-cardiac arrest setting.

## 1 Introduction

Targeted temperature management (TTM) has been widely regarded as a promising therapy in the resuscitation of cardiac arrest patients primarily because it is believed to improve neurological function [1, 2, 3, 4]. However, the benefits of TTM have not been consistently observed across trials [5, 6]. Moreover, TTM is resource intensive and can lead to serious adverse effects [7].

The rationale for TTM is based on extensive preclinical studies demonstrating attenuation of biologic mechanisms in hypoxic-ischemic brain injury and higher levels of neurological function in animals receiving early cooling after experimentally induced arrest. The inconclusive and inconsistent results observed in clinical trials reflect heterogeneous treatment effects [8], in which differentiated responses to TTM would be contingent on unaccounted biological variance in the cardiac arrest population. Deeper insights into this heterogeneity could lay the foundation for differentiated therapeutic strategies that align more closely with characteristics of individual patients or subsets of the latter, generating more reliable treatment responses.

The ability to predict postcardiac arrest outcomes has proven remarkably challenging, in part because of a complex pathophysiology involving ischemia-reperfusion injury in multiple tissues and organ systems. Prognostic scores have been proposed based on statistical models combining different variables to generate a probability of death or neurological recovery following cardiac arrest. Ostensibly designed to assist clinicians in optimizing treatment decisions, these scores are typically used as benchmarks but are rarely implemented in clinical practice. Traditional modeling approaches may not account for the heterogeneity and dynamic complexity of cardiac patients, leading to risk estimates which may not be relevant in specific clinical settings or for individual patient characteristics.

Recently, there has been growing interest in the possibility of leveraging machine learning (ML) to support classification and prediction tasks after cardiac arrest. We demonstrated that ML applied to physiological data recorded in the early phase after CA resuscitation could accurately and reliably predict postcardiac arrest outcomes [9]. Here we build on this prior work to predict outcomes associated with TTM.

The aims of this study were twofold. First, to predict the discharge outcome of postcardiac arrest patients receiving TTM by training a ML model with physiological data and TTM treatment data. Second, to identify specific features that are associated with favorable outcome following TTM. We hypothesized that patients most likely to benefit from TTM would have a unique data signature. Such a model could enable greater precision in the selection of potential TTM candidates, establishing a baseline for personalized cardiac arrest treatment, and enable cohort enrichment for future clinical trials.

## 2 Methods

### Patient Selection

Data were extracted from the multicenter Philips eICU - Clinical Research Database [10]. Patients were included if they were adults admitted to ICU after cardiac arrest. Since TTM is not consistently recorded in eICU, we analyzed temperature time series recorded in the first 24 hours after ICU admission. We designated as TTM any instance of a patient whose body temperature decreased to < 36°C after admission and remained below that threshold for > 12h in the first 24h after admission. All patients with potentially erroneous recordings, as signified by infeasible body temperatures less than 20°C included in the database, were excluded from the study. A total of 444 patients meeting these criteria were identified. Among the latter, neurological outcome data was available for 310 patients (Fig. 1).

**Figure 1:**
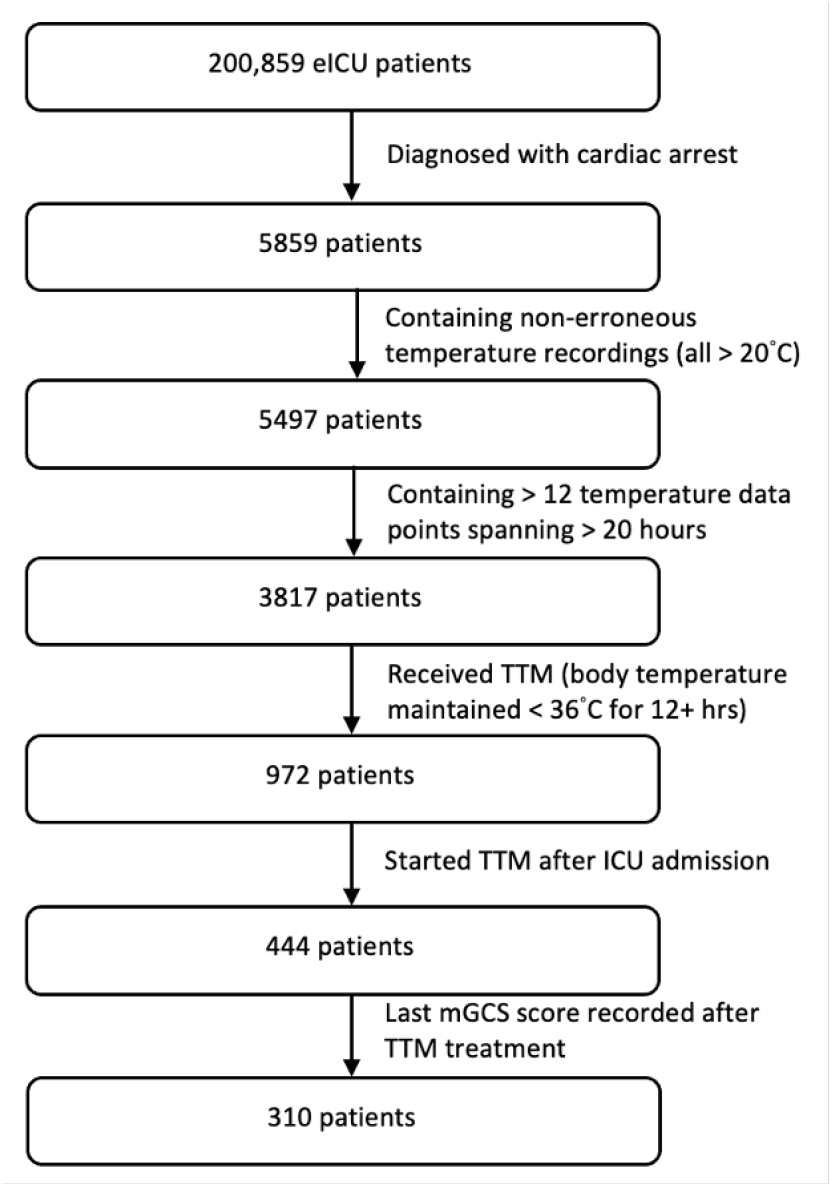
Patient selection process. From all the patients from the Philips eICU database, cardiac arrest patients who received TTM were identified. Only patients whose records included sufficient datapoints were included.

### Dataset Curation

The primary task was to predict post-TTM outcome at the time of discharge from ICU. The two outcome variables were survival at discharge and neurological function at discharge. Neurological function was defined using the motor subscore of the Glasgow Coma Scale (mGCS). A mGCS of 6 signifies a patient who is awake and able to follow verbal commands, while an mGCS < 6 implies greater neurological impairment. We used the mGCS as the neurological outcome variable, since more detailed assessments were not available in eICU; the rationale for this approach has been described elsewhere [9].

Variables used to predict outcome were extracted from clinical data collected from the time of ICU admission to the time when TTM was initiated or 12 hours post admission, whichever came first. This interval was selected as the window during which treatment decision-making was most likely occurring. Patient data included demographics; medical history; laboratory test results for the first 12 hours of ICU admission or up to the start of TTM treatment, whichever came first; type of initial heart rhythm detected; and length of CPR administered, if the CA was witnessed (Table 2).

For physiologic time series and laboratory data, we derived mean, standard deviation, minimum, maximum, first, and last values recorded. Only lab tests for which > 50% of the patients had data were used. After removing less frequent lab tests, we employed a random forest-based imputation using the randomForestSRC package for any missing values [11].

To help with identification of the most important features, we also implemented forward selection with a relative likelihood threshold of 0.5. Alias features are computed and removed to eliminate highly correlated variables. AIC values are computed from the GLM models. If their relatively likelihood, defined as their exponential halved difference, exceeds the 0.5 threshold, the feature is incorporated for models predicting mortality and neurological function.

### Modeling and Outcome Variable

We employed three modeling methods: generalized linear modeling (GLM), random forest (RF), and gradient boosting (XG) [12, 11, 13]. Hyperparameters used were the default for each package to allow for equal comparison across the different models. Models were 10-fold cross-validated and resampled 10 times once again. The models were used to assess patient mortality status at hospital discharge and neurological status following TTM treatment.

### Model Performance Evaluation

To evaluate performance of the models, we computed the Area under the Receiver Operator Characteristic (AUROC), calibration, sensitivity, specificity, accuracy, precision, and F1 with the EvaluationMeasures package for each iteration of training and computed the mean across all 10 folds [14].

The models output a classification on whether individual patients will survive and whether their mGCS will achieve a normal score of 6. To interpret which features are more indicative of the outcome, we computed the mean Shapley Additive Explanations (SHAP) values with the SHAPforxgboost package for high-predicting features recorded prior to the start of TTM [15]. Positive SHAP values indicate that a positive value of the feature helped to predict a positive outcome (survival for mortality model and mGCS of 6 for neurological outcome model) while negative SHAP values indicate that a positive feature value is correlated with a negative outcome (death for mortality model and mGCS less than 6 for neurological outcome model).

We also assessed the beta coefficients output from the GLMs with the caret package for analyzing effects of each feature on the GLM predictions [16]. A greater magnitude of the coefficient indicates greater weight of that feature. A positive value coefficient indicates a direct correlation between higher values of the feature and survival/normal motor scores while negative coefficients represent direct correlation between higher values of the features and death/lower motor scores.

## 3 Results

### Patient and Feature Selection

Among the 200,859 patients from the eICU database, 5859 were diagnosed with cardiac arrest (CA) (Table 1). As outlined above, patients with outlier body temperature recordings (less than 20°C) were removed, leaving 5497 patients. Of those, 3817 had temperature recordings, with at least 12 data points spanning the first 20 hours following admission. Of the latter, 972 patients had body temperatures which remained continuously *<* 36°C for *≥* 12 hours in the first 24 hours following admission – these were designated as having received TTM. To account for consistent TTM administered, we limited patients to those receiving TTM after ICU admission, for a total of 444 TTM patients. Of those, 310 had neurological outcome labels available, who were included for this study.

After capturing our patient population, we integrated forward feature selection to limit the number of features the models should be trained with, given the large number of features we had in the initial feature space (331). Following forward selection with a 0.5 threshold, we were left with 35 features predicting mortality and 40 features for predicting mGCS.

### Mortality Outcome Prediction

To predict mortality upon discharge, we developed three models, employing the GLM, RF, and XG methods. The best performing model, GLM, achieved an AUROC of 0.86 ± 0.04, accuracy 0.76 ± 0.04, sensitivity 0.75 ± 0.09, specificity 0.77 ± 0.07, precision 0.83 ± 0.04, and *F*_1_ 0.78 ± 0.05 across 10 cross-validations (Fig. 2).

**Figure 2:**
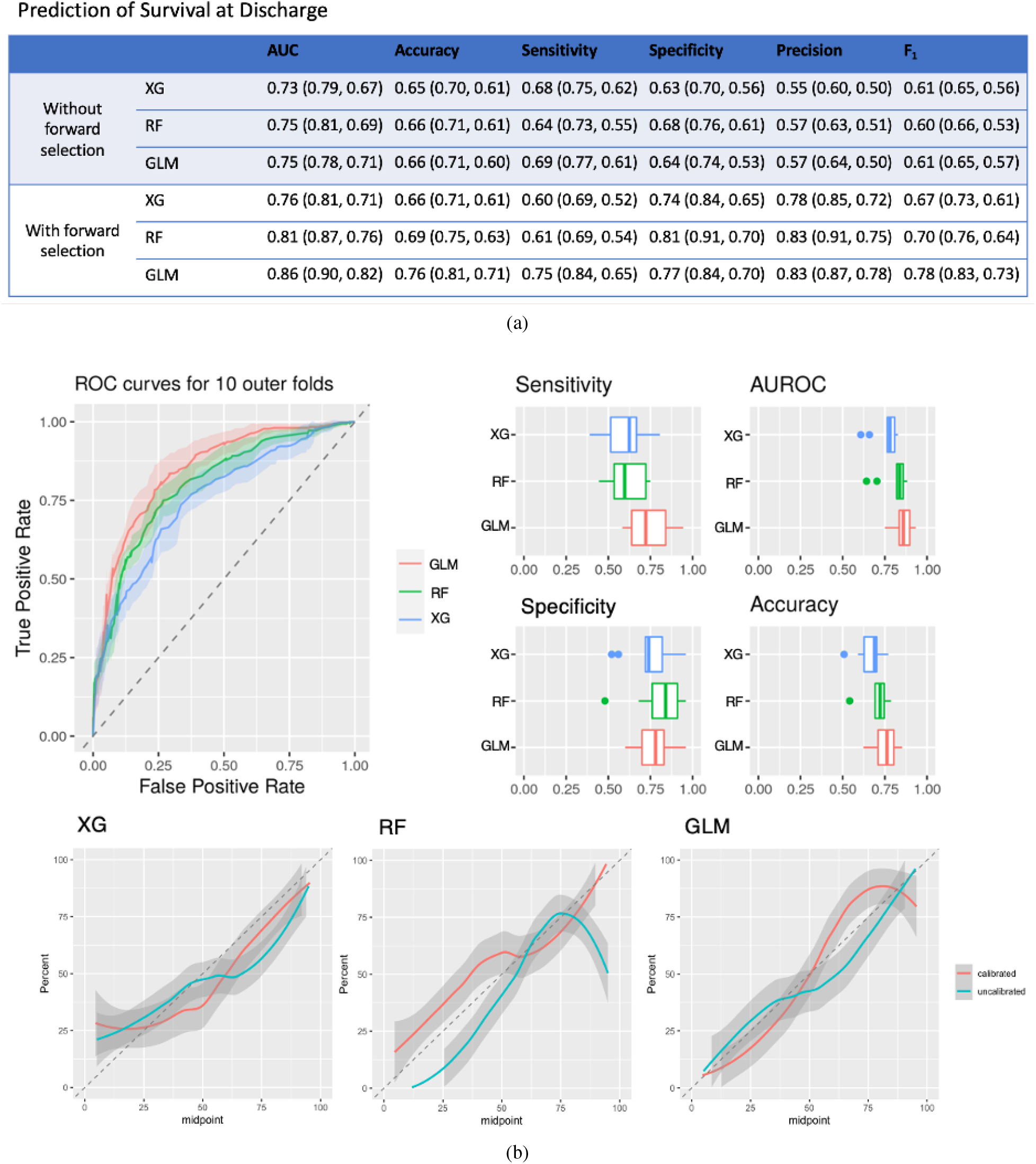
Modeling results predicting mortality upon discharge. After the initial round of modeling, the top features were selected via forward selection with a threshold of 0.5, and models were developed again with the most predictive features, resulting in increased model performance across all three model types (a). For the models with forward selection features, AUROC and calibration curves (with shaded regions representing ± 1 SD), as well as sensitivity, specificity, and accuracy ranges are shown (b).

For neurological function prediction, GLM performed slightly better than RF, with an AUROC of 0.75 ± 0.03, accuracy 0.66 ± 0.05, sensitivity 0.69 ± 0.08, specificity 0.64 ± 0.10, precision 0.57 ± 0.07, and F1 0.61 ± 0.04. Similar to mortality predictions, top predictive features were identified via forward selection with 0.5 threshold, and models improved significantly, with GLM achieving an AUROC of 0.85 ± 0.03, accuracy 0.77 ± 0.05, sensitivity 0.71 ± 0.10, specificity 0.80 ± 0.12, precision 0.73 ± 0.08, and *F*_1_ 0.71 ± 0.04 across 10 cross-validations(Fig. 3).

**Figure 3:**
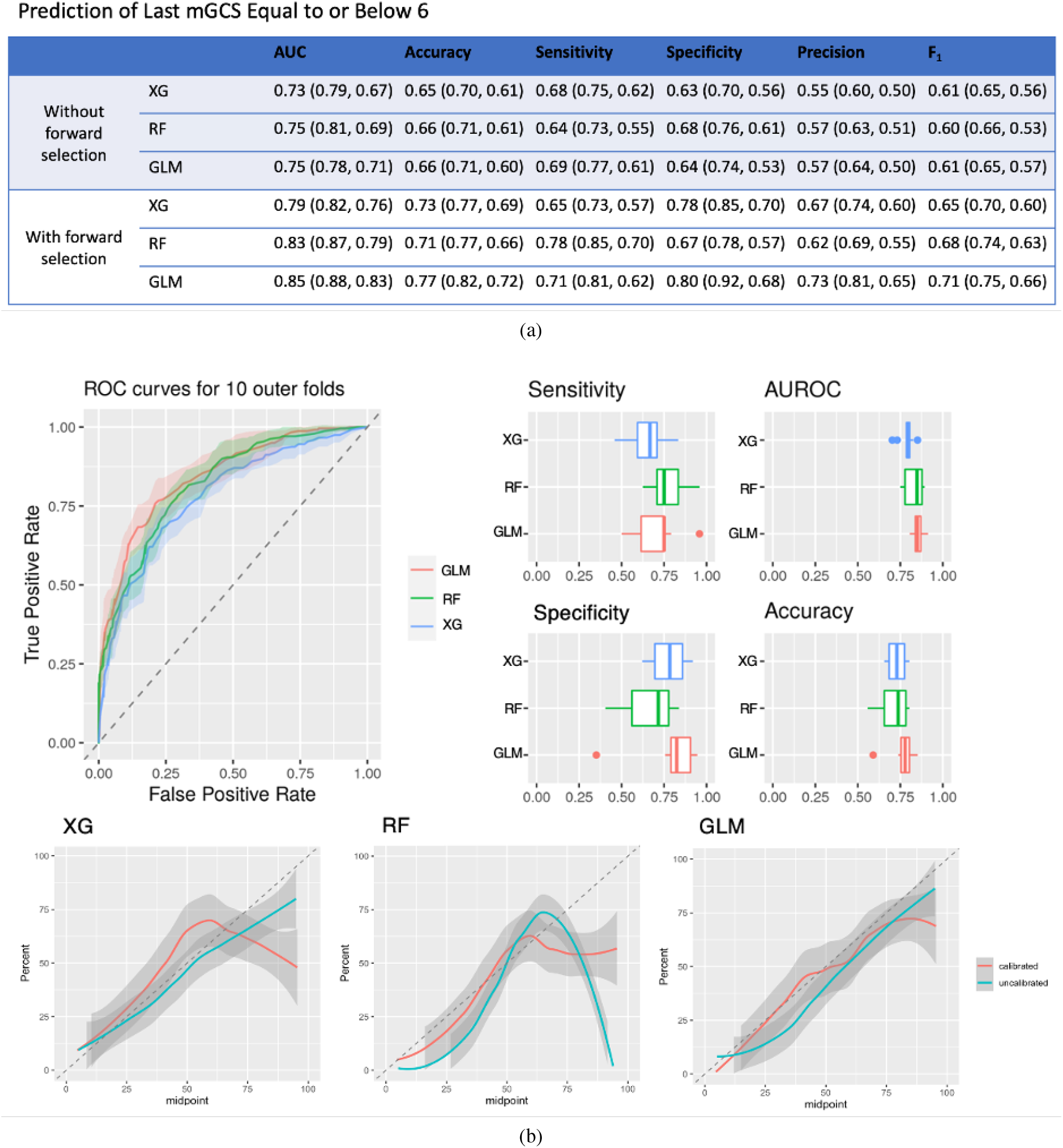
Modeling results predicting last mGCS. With a threshold of 0.5, top features were identified via forward selection, and new models were trained, which resulted in improved modeling results (a). ROC curves and calibration plots with shaded regions of ± 1 SD and sensitivity, AUROC, specific, and accuracy ranges are shown (b).

Some of the highest scoring SHAP (most informative) features included the last pH recording, last platelet count data point, last white blood cell (WBC) count, and first lactate recording, where first refers to the first recording captured for the patient after hospital admission and last for the final value recorded prior to the start of TTM. Higher values of the last pH and platelet count had a high contribution towards predicting survival, whereas higher values of first lactate value and last WBC count had a low contribution towards survival. However, lower values of last pH and platelet count had a low contribution towards predicting death while lower values of first lactate level and last WBC count recorded had higher contributions towards predicting death (Fig. 4a). The SHAP values also match the beta coefficient calculated for each feature in the GLM, in which higher values of last pH and last platelet counts are both predictive of survival and higher values of first lactate and last WBC count are predictive of expiration.

**Figure 4:**
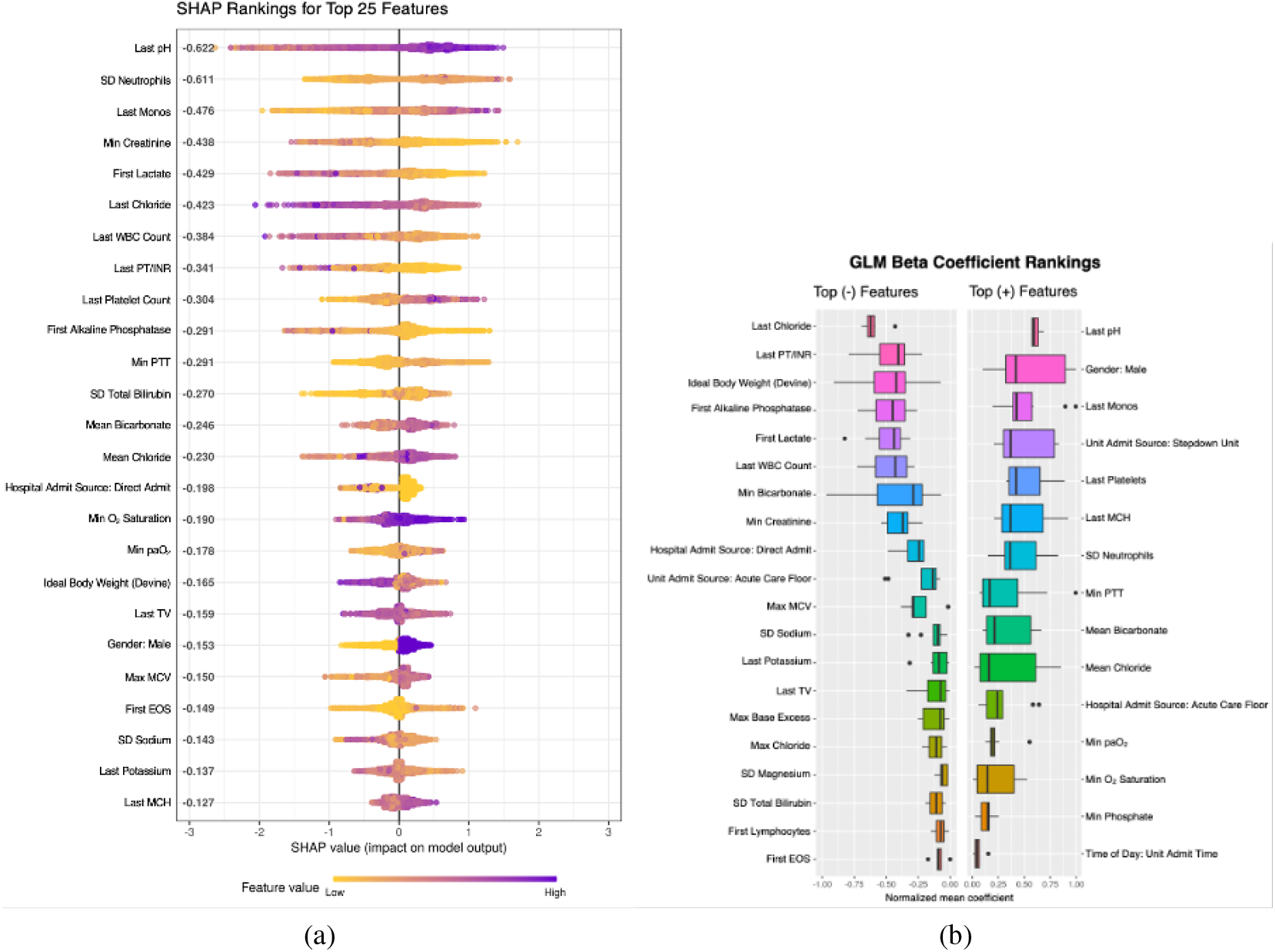
Top predictive features predicting mortality based on SHAP values and GLM beta coefficients. Mean SHAP values are shown to the right of each feature name. Positive SHAP values indicate survival while negative value indicate death at hospital discharge. Purple/yellow bars are representative of higher/lower values of the feature (a). Features ranked by beta coefficients in the GLM are predictive of survival for positive normalized mean coefficients and death for negative normalized mean coefficients (b).

### Neurological Outcome Prediction

We also used SHAP to rank features for predicting neurological function. We observed some similar top predictive features such as last pH measured, which also demonstrates positive correlation with mGCS. Other features include minimum monocyte count, which also highly contributes towards mGCS of 6 predictions, and last FiO2 levels and first base excess lab test, both of which highly contribute towards patient outcomes with mGCS less than 6 (Fig. 5a). Similar trends are shown with the GLM beta coefficient rankings, where the last pH and min monocyte count are both positive predictive features and first base excess value and last FiO2 level are negative predictive features (Fig. 5b).

**Figure 5:**
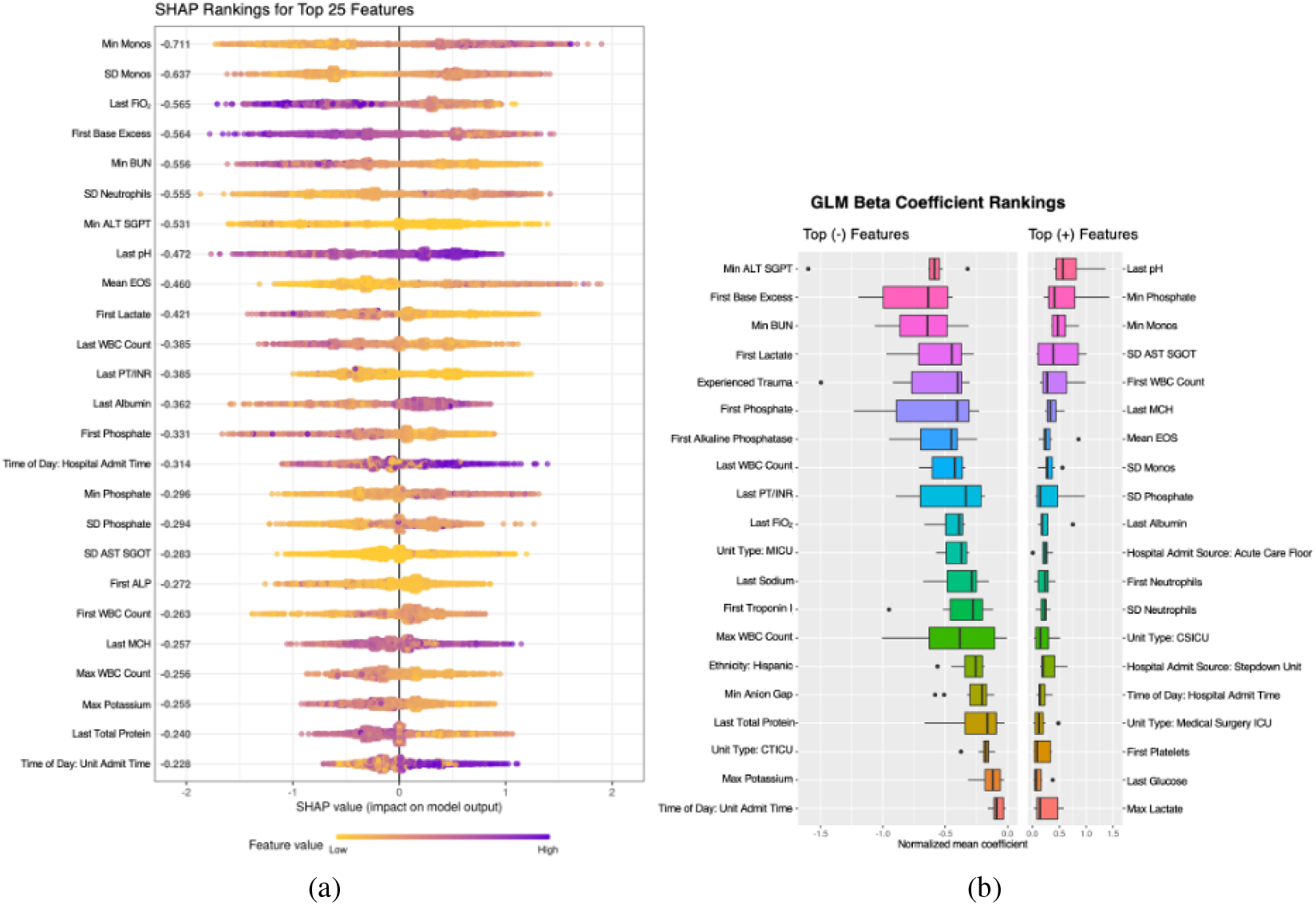
Top predictive features predicting mGCS of 6 or mGCS < 6 based on SHAP values and GLM beta coefficients. Mean SHAP values are shown to the right of each feature name. Positive SHAP values represent the last recorded mGCS after TTM treatment with a value of 6, and negative SHAP values represent the last recorded mGCS after TTM treatment with a value of less than 6; purple bars indicate high values of the feature, and yellow indicate low feature values (a). Coefficients are ranked based on beta coefficients from the GLM, with positive normalized mean coefficients demonstrating mGCS of 6 and negative demonstrating mGCS < 6 (b).

## 4 Discussion

In the analysis of this postcardiac arrest patient dataset, we employed supervised ML classifiers that accurately predict post-TTM outcomes. These interpretable models identified several previously unreported features predictive of favorable outcome after TTM. Data collected from routine laboratory tests proved to be most predictive of both mortality and mGCS outcomes. We discovered that higher values of last pH recording and platelet count positively predict survival while higher values of first lactate label and last WBC count are predictive of death. Higher pH measured and minimum monocyte count are predictive of favorable neurological function while higher base excess values and FiO2 levels predict worse neurological function.

Overall, our study provides insight into physiologic and laboratory features that impact outcomes in cardiac arrest patients receiving TTM. By limiting the dataset to incorporate only the most predictive features via forward selection, our models were able to better capture the correlations between individual features and results, as indicated by the increased performance of models. To the extent of our knowledge, no other study has been completed that identifies features predicting outcomes of TTM treatment.

As findings are based on a large dataset with multiple institutions, results from this study can be incorporated into cardiac arrest care protocol across different hospitals. Leveraging clinical data collected regularly during the first hours of ICU admission, these models can be used in conjunction with current treatment planning strategies to optimize care for post-cardiac arrest patients. Ideally, TTM should be administered soon after admission to the hospital, but any data collected prior to treatment can provide insight into whether a CA patient is a suitable candidate for TTM [17, 18].

There are several limitations in this study. The cardiac arrest patients in the dataset were not designated as recipients of TTM treatment or not. This resulted in the need to develop an algorithm to check for temperature data points to determine whether TTM was administered. Some TTM patient temperature data fluctuate quite significantly, and these patients would have been excluded from the study had their temperatures oscillated above and below 36°C. TTM was also not administered in the same manner across all patients as the eICU dataset incorporated patients across various hospitals. Furthermore, we were unable to externally validate our result and confirm generalizability due to lack of access to other additional cardiac arrest datasets, but cross-validation was implemented to prevent overfitting. As this was a retrospective study, some labels and features that could potentially provide useful insight into patient outcome were not available, such as Cerebral Performance Category (CPC) scores that could be used to evaluate neurological outcome.

In the future, we hope to investigate how different parts of the TTM, such as rates of cooling, rewarming, duration of cooling, and offset of TTM administration, could affect physiological outcomes. Additionally, we will optimize models with further hyperparameter tuning to evaluate the best prediction outcomes that GLMs can achieve. Once we gain access to other datasets and can verify that models robustly predict mortality and mGCS outcomes across different patient data, we hope to develop a clinical study specifically for cardiac arrest patients who are potentially candidates of TTM. This will enable us to gather more clinically predictive features and evaluation labels, such as CPC, serum biomarkers, and imaging and EEG data. Such a prospective study will help us better understand how physicians can utilize these models for predictions in real time and guide their treatment plans.

Overall, these models hold potential to aiding personalized treatment planning of cardiac arrest patients, providing predictions of whether a post-cardiac arrest patient would survive and if they can retain normal motor function following TTM. Given a patient’s physiological and lab data during their first hours of admission to the ICU, ML models can provide interpretable results regarding post-cardiac arrest patient prognostics.

## Data Availability

All data produced in the present study are
available upon reasonable request to the authors.

